# The impact of COVID-19 pandemic on health service utilization among patients with chronic non-communicable diseases in Ghana

**DOI:** 10.64898/2026.09.03.26362130

**Authors:** Elliot Koranteng Tannor, Sam Newton, Reinhard Busse, Sylvanus Gatorwu, Joshua Kwasi Safo, Richmond Adjei, John Amuasi, Wilm Quentin

## Abstract

**Background:** The COVID-19 pandemic significantly disrupted health systems globally and in sub-Saharan Africa (SSA) leading to reduction of health service utilization (HSU). Patients with non-communicable diseases (NCDs) had increased morbidity and mortality during the COVID-19 pandemic. We sought to determine the changes and impact of the COVID-19 pandemic on HSU among patients with NCDs comparing an epicentre to a non-epicentre region in Ghana.

**Methods:** We conducted a cross-sectional observational study of patients with NCD in the Ashanti and Northern regions in Ghana from 14^th^ April 2024 to 31^st^ July 2024. We distributed questionnaires to patients with NCD in selected facilities in the Northern and Ashanti regions of Ghana. The Andersen’s Behavioural Model was used as the conceptual framework to determine the factors affecting HSU. Multiple logistic regression was used to identify independent variables predicting reduction in HSU. P value of less than 0.05 was considered statistically significant.

**Results:** The study included 844 participants with NCDs equally sampled in the Ashanti and Northern regions of Ghana. The mean age of respondents was 61.4±11.6 years with 569(67%) females. Almost half 367 (43.5%) of all respondents reported COVID-19 led to a decrease in their HSU and over a third of respondents, 303(35.9%) missed their hospital appointments. The proportion of respondents reporting a decrease in HSU was significantly higher in the Northern region 276 (65.4%) than the Ashanti region 91 (21.6%), [p<0.001]. Reduction in HSU was associated with increasing age (aOR=0.97, 0.96 - 0.99, p<0.001), Northern region residence (aOR=9.60 CI 6.43 - 14.32, p<0.001), higher household income (aOR=1.36 CI 1.15-1.60, p<0.001), longer duration of NCD (aOR=1.03, CI 1.01 - 1.06, p=0.033) and possession of valid NHIS card (aOR=7.43, CI 2.44 - 22.57, p <0.001).

**Conclusion:** The COVID-19 pandemic led to reduction in HSU in Ghana. There was a significant reduction in NCDs in a non-epicentre region (Northern region) compared to the Ashanti region which was an epicenter region. Hence patients in non-epicentres should also be prioritized in pandemic situations.

## Introduction

The COVID-19 pandemic has significantly disrupted health systems globally in low- and lower-middle income countries (LLMICs), particularly in sub-Saharan Africa (SSA) [1–4] leading to reduction of health service utilization (HSU)[5]. During the COVID-19 pandemic, health services were globally disrupted in 94% of countries according to the World Health Organization (WHO)[6]. The incidence and mortality from COVID-19 infection was reportedly lower in SSA as compared to other continents[7, 8] but SSA still experienced significant disruptions and reduction in HSU[9].

The disruptions in health service delivery led to a decrease in the provision of essential healthcare services in the management of communicable diseases, injuries and non-communicable diseases (NCDs). Non-communicable diseases (NCDs), including cardiovascular diseases, diabetes, chronic respiratory diseases, and cancers, pose significant challenges to health systems worldwide[10]. According to the WHO, NCDs are responsible for about 41 million deaths, equivalent to 71% of all deaths globally, and account for 85% of deaths between 30-69 years in LMICs [10]. NCDs also increased the risk of severe infections, morbidity, and mortality in patients with COVID-19 during the pandemic [11]. Among NCDs, patients with diabetes and hypertension were the most reported to be impacted significantly with increased morbidity and mortality during the COVID-19 pandemic [12].

Health Service Utilization is the process of seeking professional healthcare and submitting one’s self to the application of the regular health services with the purpose to prevent or treat health problems[13]. HSU is a reflection of the health seeking behaviour of a people, which is dependent on demographic, socioeconomic status, geographical location and the severity of the health condition [14].

HSU is an important health outcome indicator and also an independent variable that predicts other health outcomes such as perceived health status, evaluated health status and satisfaction of the healthcare consumer [15]. The Andersen’s Behavioural Model[16] helps to identify the determinants of HSU including both individual and contextual factors. Individual factors are grouped into predisposing factors, enabling factors and need factors in the context of their families and communities [15–17].

COVID-19 pandemic has been shown to lead to reduction in HSU in SSA. A recent scoping review in 39 countries in SSA showed a decrease in HSU amongst 84.4% the 262 studies included, with a median decrease in HSU of 35.6%. The patient group with the largest percentage decrease in HSU was those with cardiovascular diseases, with a median decrease of 68% in HSU in SSA [9]. Effective management of NCDs is dependent on access to health services, diagnostics and medications. However, during the COVID-19 pandemic, there were several barriers including lockdown, fear of infections, transport restrictions and reallocation of staff and resources towards pandemic control [18].

The reports of COVID-19 cases during the pandemic in Ghana, according to the Ghana Health Service(GHS) showed that the highest number of cases were in the Greater Accra and Ashanti regions with the least cases in the Northern regions[19]. In Ghana, the Government of Ghana and Ministry of Health implemented policies to reduce the spread of the infection and mitigate the impact of the COVID-19 pandemic of health service delivery but data on the direct effect of the pandemic on NCDs is lacking. There were reported reduction in hypertension claims by 20% according to the National Health Insurance Authority during the pandemic in 2020 but there was a reported increase in severity of hypertension as measured by the number of hypertension episodes with an increase of 18.6% in the cost per hypertension per year [20].

The effect of the COVID-19 pandemic on HSU among patients with NCDs in Ghana has not been studied. We sought to determine the changes and impact of the COVID-19 pandemic on HSU comparing HSU among patients with non-communicable diseases (NCD) in one of the regional epicentres to a non-epicentre region during the pandemic in Ghana using the Andersen’s Behavioural Model as the conceptual framework.

## Methods

We conducted a cross-sectional observational study of patients with chronic non-communicable diseases (hypertension and diabetes mellitus) in Ghana. The study was conducted in selected health facilities in the Ashanti and the Northern regions of Ghana.

### Ashanti Region

The Ashanti Region is one of the country’s sixteen administrative regions. The Ashanti Region covers an area of approximately 24,389 square kilometers, making it the third-largest region in Ghana by land area. Kumasi, the regional capital, serves as a major urban and economic hub. As of the 2021 Population and Housing Census, the Ashanti Region was the most populous region in Ghana, with approximately 5.4 million inhabitants accounting for about 18.9% of the national population. Kumasi, the regional capital, is the second-largest city in Ghana with a significant urban centre [21]. The Ashanti region (22,640 cases) was second to Greater Accra (97,680 cases) as the highest number of COVID-19 cases during the pandemic according to the Ghana Health Services (GHS) as at April 2024[19].

### Northern Region

The Northern Region is located in the northern part of Ghana, is known for its vast land area and diverse ethnic groups. The Northern Region covers an area of approximately 70,384 square kilometers, making it the largest region in Ghana by land area. Tamale, the regional capital, is a growing urban centre and a hub for commerce and education. As of the 2021 Population and Housing Census, the Northern Region had a population of approximately 2.3 million, accounting for about 7.9% of Ghana’s total population. The population is predominantly rural, with agriculture being the primary livelihood for most inhabitants [21]. The number of recorded COVID-19 cases during the pandemic in the Northern region was 1930 cases as at April 2024 [19].

### Selected study health facilities

Three health facilities were selected purposively to be included in the study in each of the two regions. The selection was based on comparable out-patient clinic volumes according to the DHIMS-2 database. The primary, secondary and tertiary facilities were the Ejisu Juaben Health centre, Kumasi South Hospital and Komfo Anokye Teaching Hospital respectively. The primary, secondary and tertiary hospitals selected in the Northern region were Savelugu District Hospital, Tamale Central Hospital and Tamale Teaching Hospital (TTH) respectively (Figure 1).

**Figure 1.**
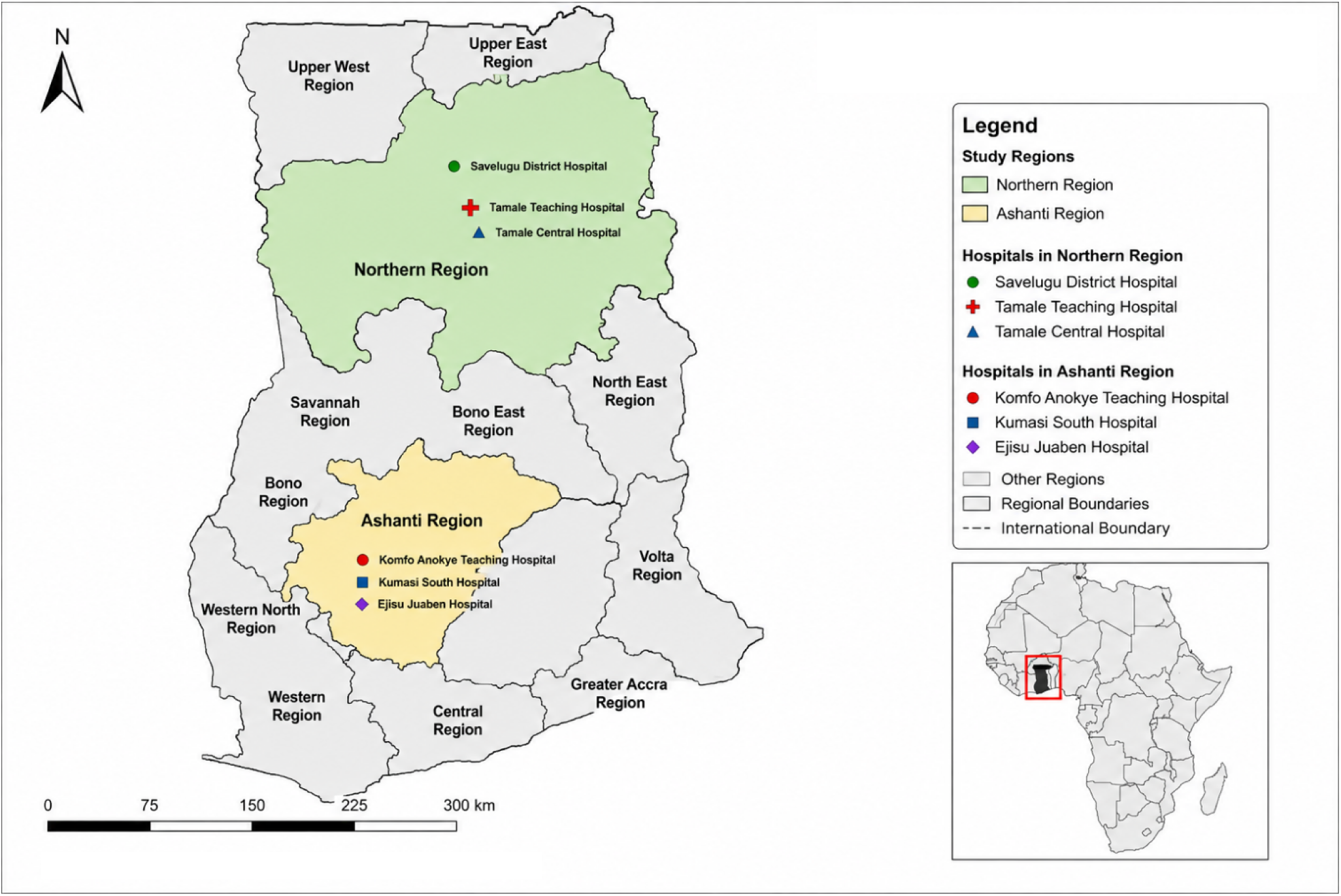
Map of Ghana showing the study regions and study sites

### Sample size

Using logistic regression for observational studies, sample size was determined based on the number of independent variables in the model for the multivariable analysis [22]. Calculation was done based on an expected number of independent variables of ten from the Andersen’s Behavioral Model conceptual framework for change in HSU as the primary outcome[17]. A total of 844 participants were recruited for the study.

### Study population

The study population included patients aged 18 years and above with hypertension and/or diabetes mellitus enrolled in chronic care clinic for at least 12 months before the COVID-19 pandemic who gave a written informed consent. Data was collected from 14^th^ April 2024 to 31^st^ July 2024.

Participants were excluded if they failed to recall past events or had self-reported history of dementia.

### Study Procedure

Participant with NCDs attending the selected facilities for routine care who gave an informed consent were recruited into the study. Participants were given a pre-tested questionnaire to ascertain their experience before and during the COVID-19 pandemic in Ghana. The questionnaire captured their demographics, socioeconomic data, health service needs and HSU before and during the pandemic, reasons for any change in HSU, the impact of the COVID-19 pandemic and change in HSU on their perceived, evaluated health and their satisfaction with health services. Participants were requested to complete the questionnaire by themselves but there was also translation to the local language for participants who could not complete the questionnaire in English. The data was collected onto an online application EpiCollect 5[23] from the various centres on the same electronic platform.

The Andersen’s Behavioural Model [17] was used as the conceptual framework to determine the factors determining change in HSU and impact of COVID-19 on HSU. This model was chosen because it is comprehensive and has a structural approach with system-level factors and individual level factors and the most utilized in measuring HSU. Predisposing factors include age, gender, ethnicity and marital status; enabling factors included were educational level, income status, employment status, household size and availability of health insurance; Need factors refers to need for health service and included disease severity, duration of chronic illness and presence of acute disease. The study outcomes were perceived health status, evaluated health status, patient satisfaction with health service (Figure 2).

**Figure 2.**
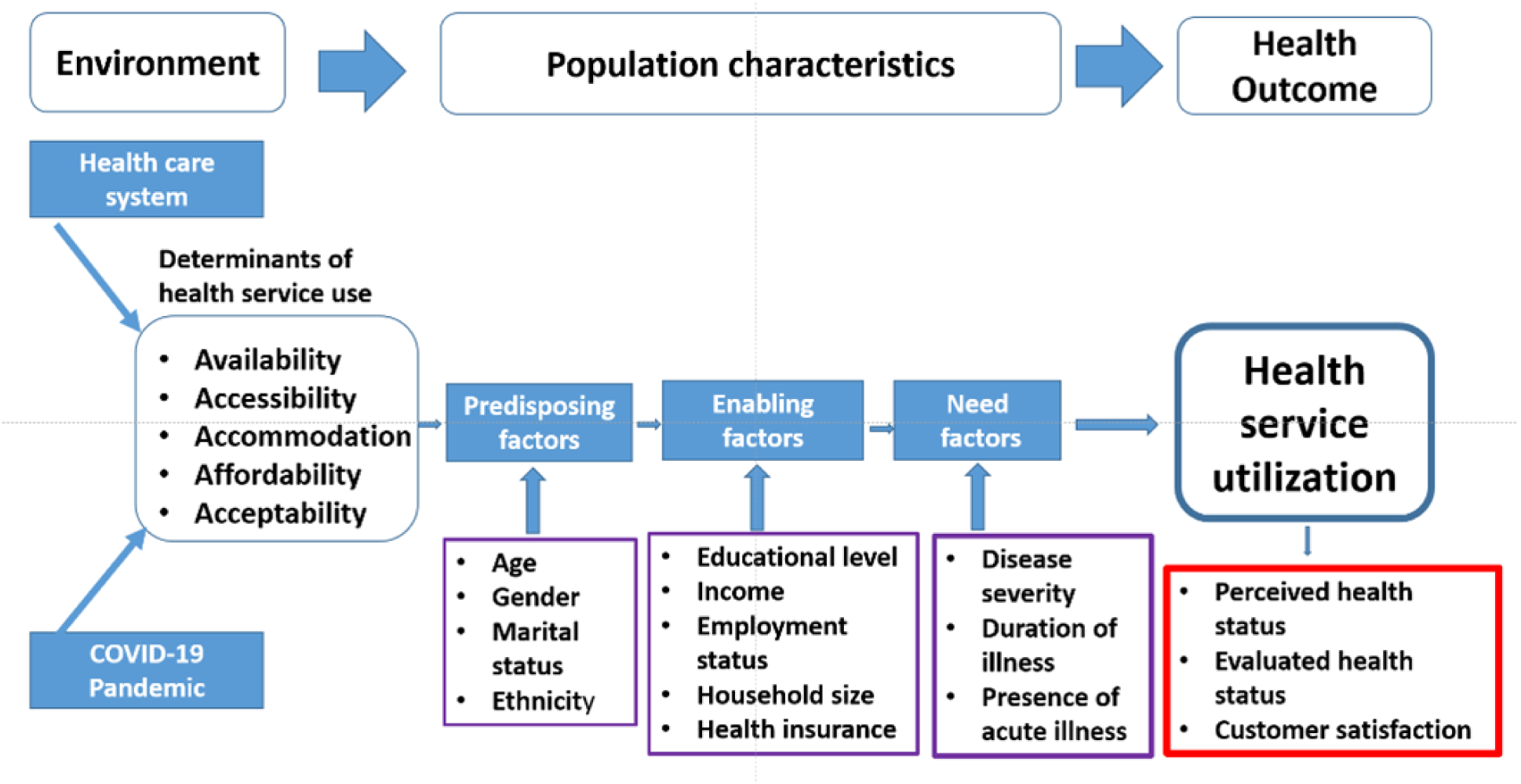
The conceptual model of the determinants of HSU combining Anderson’s Behavioral Model of HSU[17] and Penchansky and Thomas’ Access to health model [24] (Source: Author’s Construct 2023)

Patient satisfaction with health service was assessed with the Patient Satisfaction Questionnaire (PSQ) short form questionnaire and categorized as satisfied or dissatisfied. A mean score of >3 was considered satisfied and those <3 categorized as dissatisfied for subscales including General satisfaction (2), Technical quality (4), Interpersonal manner (2), Communication (2), Financial aspects (2), Time spent with doctor (2), and Accessibility and convenience (4) [25].

## Statistical analysis

Categorical variables were summarized as proportions and percentages. Continuous variables were summarized as means and standard deviations when parametric and median with interquartile range when non-parametric. Student t-test and Wilcoxon signed rank test were used to test continuous variables when parametric and non-parametric respectively, to determine the factors that are associated with a reduction in HSU. Multiple logistic regression was used to identify independent variables predicting reduction in HSU in the Andersen’s Behavioral Model. P value of less than 0.05 was considered statistically significant.

## Ethical Considerations

Ethical clearance was granted from the Ghana Health Service Ethics Review Committee with approval number GHS-ERC/010/01/24, the Committee on Human Research and Publication Ethics (CHRPE) of KNUST with approval number CHRPE/AP/1045/23 and the institutional review board of the Komfo Anokye Teaching hospital with approval number KATH/IRB/AP/009/24 before the conduct of the study.

## Results

### Demographic characteristics of respondents in the Survey

The study included 844 participants with chronic non-communicable disease (hypertension and/or diabetes) equally sampled in the Ashanti and Northern regions of Ghana. The mean age of respondents was 61.4±11.6 years with 569(67%) females. The majority of respondents were married 526 (62.3%), Christians 476 (56.4%) and Akans 386 (45.6%) with 347 (41.1%) Dagombas (Table 1). The median duration of NCD diagnosis among respondents was 8 years (IQR 5-11). Almost half, 367 (43.5%) of all respondents reported that COVID-19 led to a decrease in their HSU. About a third, 303 (35.9%) of all respondents missed their medical appointments as a result of the COVID-19 pandemic. About a fifth 155 (18.4%) of respondents were acutely ill during the pandemic and 118 (14.0%) were hospitalized (Table 1).

**Table 1.** Demographic characteristic of all respondents and by region n=844.

| Variable | All<br>n=844 | Ashanti<br>(n=422) | Northern<br>(n=422) |
| --- | --- | --- | --- |
| Age (years) u±SD | 61.4 ±11.6 | 62.7±11.3 | 60.1±11.7 |
| Female Gender n (%) | 569 (67.4) | 322 (76.3) | 247(58.5) |
| Marital status n (%) |  |  |  |
| Single | 46 (5.5) | 23(5.5) | 23 (5.5) |
| Married | 526 (62.3) | 210 (49.8) | 316 (74.9%) |
| Divorced/Separated | 91 (10.8) | 69 (16.4) | 22 (5.2%) |
| Widowed | 181 (21.5) | 120 (28.4) | 61 (14.5) |
| Religion n (%) |  |  |  |
| Christian | 476 (56.4) | 400 (94.8) | 76 (18.0) |
| Muslim | 368 (43.6) | 22 (5.2) | 346 (82.0) |
| Ethnicity n (%) |  |  |  |
| Akan | 386 (45.7) | 378 (89.6) | 8 (1.9) |
| Dagomba | 347 (41.1) | 1 (0.2) | 346 (82.0) |
| Others | 111 (13.2) | 43 (10.2) | 68 (16.1) |
| Place of Residence n (%) |  |  |  |
| Rural | 127 (15.1) | 15 (3.6) | 112 (26.4) |
| Urban | 349 (41.4) | 192 (45.5) | 157 (37.2) |
| Peri-urban | 368 (43.6) | 215 (51.0) | 153 (36.3) |
| Highest level of education n (%) |  |  |  |
| None | 261 (30.9) | 81(19.2) | 180(42.7) |
| Primary | 257 (30.5) | 174 (41.2) | 83 (19.7) |
| Secondary | 196 (23.2) | 119 (28.2) | 77 (18.3) |
| Tertiary | 130 (15.4) | 48 (11.4) | 82 (19.4) |
| Employed before COVID-19 pandemic n (%) | 480 (56.9) | 239 (56.6) | 241(57.1) |
| Employed during COVID-19 pandemic n (%) | 409 (48.5) | 192 (45.5) | 217 (51.4) |
| Participants with valid National Health Insurance n (%) | 814 (96.4) | 418 (99.1) | 396 (93.8) |
| Participants with private health insurance card (%) | 43 (5.1) | 11(2.6) | 32 (7.6) |
| Total Household size n (%) |  |  |  |
| Less than 5 | 432 (51.2) | 151 (35.8) | 72(17.1) |
| 5-10 | 223 (26.4) | 247 (58.5) | 185 (43.8) |
| Greater than 10 | 189 (22.4) | 24 (5.7) | 165 (39.1) |
| Monthly net income (GHS) n (%) |  |  |  |
| Less than 500 | 353 (41.8) | 168 (39.8) | 185 (43.8) |
| 500 – 1000 | 242 (28.7) | 150 (35.6) | 92 (21.8) |
| 1000 – 2000 | 126 (14.9) | 59 (14.0) | 67 (15.9) |
| 2000 – 5000 | 106 (12.6) | 35 (8.3) | 71(16.8) |
| Greater than 5000 | 17 (2.0) | 10 (2.4) | 7 (1.7) |

u, mean; SD, standard deviation; IQR, interquartile range; NCD, non-communicable disease; GHS, Ghana cedis

### Comparison of the Ashanti to Northern regions

There was significantly a higher proportion reporting decreased HSU in the Northern region 276 (65.4%) than in the Ashanti region 91 (21.6%), [p<0.001]. Over a half of all respondents 221 (52.4%) in the Northern region missed their appointments as a result of COVID-19 pandemic as compared to 82 (19.4%) in the Ashanti region (p<0.001). There was a significantly higher portion of respondents reporting acutely ill during the COVID-19 pandemic in the Northern region 106 (25.1%) as compared to 49 (11.6%) in the Ashanti region (p<0.001), but there was no statistical difference between those who were admitted during the pandemic [55 (13.0%) vs 41 (9.7%) p=0.129] in the two regions (Table 2).

**Table 2.**
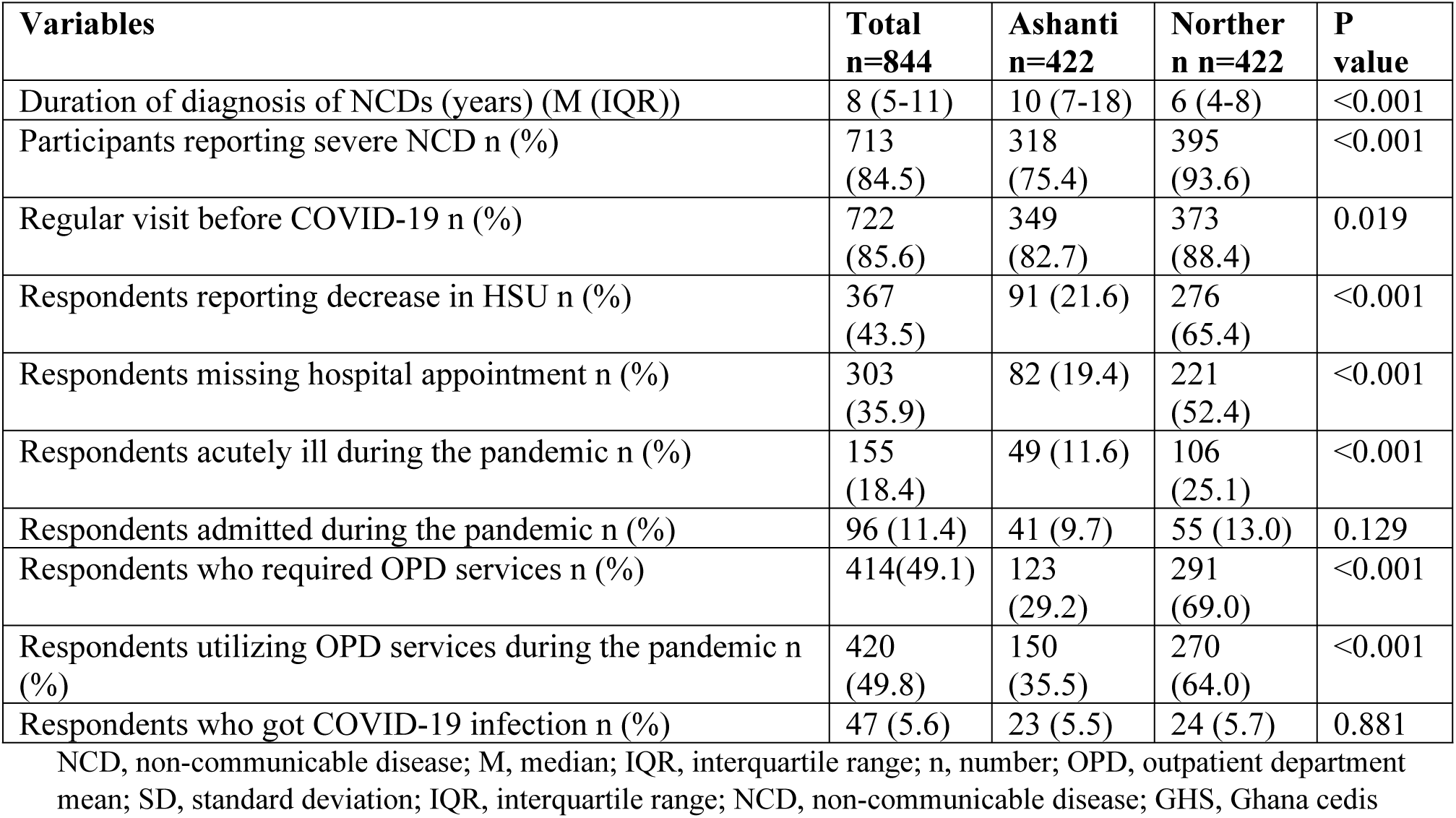
Respondents interaction with health facility in the Ashanti and Northern regions.

| Variables | Total<br>n=844 | Ashanti<br>n=422 | Norther<br>n n=422 | P<br>value |
| --- | --- | --- | --- | --- |
| Duration of diagnosis of NCDs (years) (M (IQR)) | 8 (5-11) | 10 (7-18) | 6 (4-8) | <0.001 |
| Participants reporting severe NCD n (%) | 713<br>(84.5) | 318<br>(75.4) | 395<br>(93.6) | <0.001 |
| Regular visit before COVID-19 n (%) | 722<br>(85.6) | 349<br>(82.7) | 373<br>(88.4) | 0.019 |
| Respondents reporting decrease in HSU n (%) | 367<br>(43.5) | 91 (21.6) | 276<br>(65.4) | <0.001 |
| Respondents missing hospital appointment n (%) | 303<br>(35.9) | 82 (19.4) | 221<br>(52.4) | <0.001 |
| Respondents acutely ill during the pandemic n (%) | 155<br>(18.4) | 49 (11.6) | 106<br>(25.1) | <0.001 |
| Respondents admitted during the pandemic n (%) | 96 (11.4) | 41 (9.7) | 55 (13.0) | 0.129 |
| Respondents who required OPD services n (%) | 414(49.1) | 123<br>(29.2) | 291<br>(69.0) | <0.001 |
| Respondents utilizing OPD services during the pandemic n (%) | 420<br>(49.8) | 150<br>(35.5) | 270<br>(64.0) | <0.001 |
| Respondents who got COVID-19 infection n (%) | 47 (5.6) | 23 (5.5) | 24 (5.7) | 0.881 |

NCD, non-communicable disease; M, median; IQR, interquartile range; n, number; OPD, outpatient department mean; SD, standard deviation; IQR, interquartile range; NCD, non-communicable disease; GHS, Ghana cedis

### Outcomes of health service utilization

Over a third, 309 (36.6%) of respondents reported a poor perceived health status as a result of the COVID-19 pandemic and 186 (22.0%) reported poor evaluated health status in terms of poor sugar/blood pressure control during the pandemic. Respondents in the Ashanti region reported a higher 43(10.2%) worsened evaluated health status as compared to 20 (4.7%) in the Northern region (p=0.003). Hospitalization was reported in 57(13.5%) of all respondents but there was no significant difference in reported hospitalization in the Ashanti and Northern regions of Ghana (p=0.691). Respondents in the Northern region reported higher satisfaction with the time spent with a doctor than the Ashanti region 67 (15.9) vs 44 (10.4%); p=0.019. There was however no statistical difference in perceived change in health status (p=0.673), general patient satisfaction (p=0.680), technical quality of health service (p=0.406), interpersonal manner (p=0.232), communication (0.154), Accessibility and convenience (p=0.251) and overall patient satisfaction (p=0.260) (Table 3).

**Table 3.** Patient reported outcomes as a result of their HSU in the Ashanti and Northern region.

| Variable | All<br>n=844 | Ashanti<br>n=422 | Northern<br>n=422 | P<br>value |
| --- | --- | --- | --- | --- |
| Poor perceived health status n (%) | 309 (36.6) | 170 (40.3) | 164 (38.9) | 0.673 |
| Hospitalization during pandemic n (%) | 118 (14.0) | 57 (13.5) | 61 (14.5) | 0.691 |
| Worsened evaluated health status n (%) | 63 (7.5) | 43 (10.2) | 20 (4.7) | 0.003 |
| Poor sugar or BP control n (%) | 186 (22.0) | 98 (23.2) | 88 (20.9) | 0.406 |
| **Patient Satisfaction** |  |  |  |  |
| General patient satisfaction n (%) | 108 (12.8) | 52 (12.3) | 56 (13.3) | 0.680 |
| Technical quality n (%) | 188 (22.3) | 89 (21.1) | 99 (23.5) | 0.408 |
| Interpersonal manner n (%) | 61 (7.2) | 26 (6.2) | 35 (8.3) | 0.232 |
| Communication n (%) | 131 (15.5) | 73 (17.3) | 58 (13.7) | 0.154 |
| Financial aspects n (%) | 51 (6.0) | 28 (6.6) | 23 (5.4) | 0.470 |
| Time spent with doctor n (%) | 111 (13.2) | 44 (10.4) | 67 (15.9) | 0.019 |
| Accessibility and convenience n (%) | 39 (4.6) | 23 (5.5) | 16 (3.8) | 0.251 |
| Overall patient satisfaction n (%) | 88 (10.4) | 39 (9.2) | 49 (11.6) | 0.260 |

BP, blood pressure; n, number

### Predictors of reduced health service utilization

Multiple logistic regression revealed that increasing age (aOR=0.97; 95% CI 0.96-0.99, p<0.001), Northern region residence (aOR=9.60; CI 6.42-14.3, p<0.001), increasing income (aOR=1.36; CI1.15-1.61, p<0.001), possession of valid national health insurance (aOR=7.43; CI 2.44 - 22.57, P<0.001) and duration of non-communicable disease (aOR=1.03; 95%CI 1.01-1.06, p=0.033) were the significant predictors of reduced HSU (Table 4).

**Table 4.**
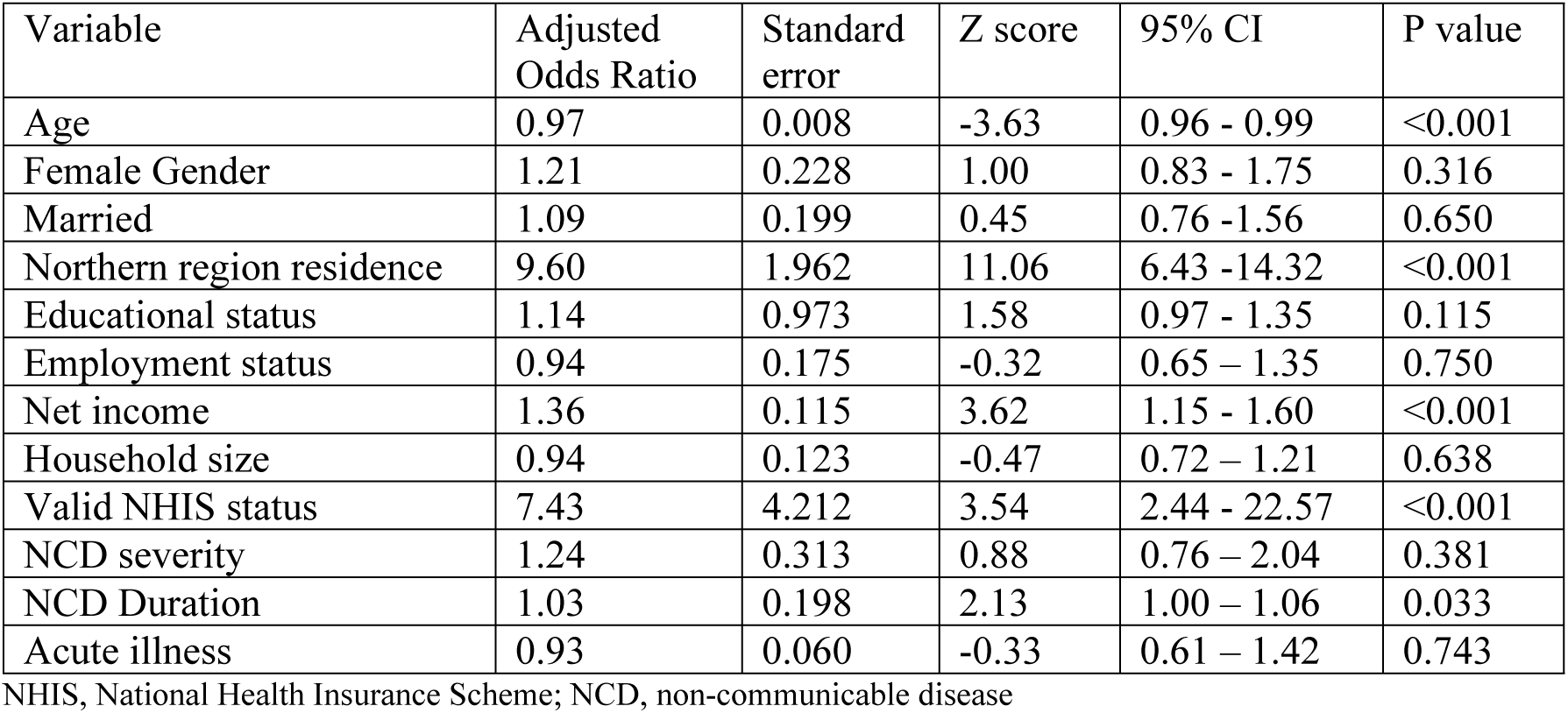
Predictors of reduced health service utilization during the COVID-19 pandemic in Ghana.

| Variable | Adjusted<br>Odds Ratio | Standard<br>error | Z score | 95% CI | P value |
| --- | --- | --- | --- | --- | --- |
| Age | 0.97 | 0.008 | -3.63 | 0.96 - 0.99 | <0.001 |
| Female Gender | 1.21 | 0.228 | 1.00 | 0.83 - 1.75 | 0.316 |
| Married | 1.09 | 0.199 | 0.45 | 0.76 - 1.56 | 0.650 |
| Northern region residence | 9.60 | 1.962 | 11.06 | 6.43 - 14.32 | <0.001 |
| Educational status | 1.14 | 0.973 | 1.58 | 0.97 - 1.35 | 0.115 |
| Employment status | 0.94 | 0.175 | -0.32 | 0.65 - 1.35 | 0.750 |
| Net income | 1.36 | 0.115 | 3.62 | 1.15 - 1.60 | <0.001 |
| Household size | 0.94 | 0.123 | -0.47 | 0.72 - 1.21 | 0.638 |
| Valid NHIS status | 7.43 | 4.212 | 3.54 | 2.44 - 22.57 | <0.001 |
| NCD severity | 1.24 | 0.313 | 0.88 | 0.76 - 2.04 | 0.381 |
| NCD Duration | 1.03 | 0.198 | 2.13 | 1.00 - 1.06 | 0.033 |
| Acute illness | 0.93 | 0.060 | -0.33 | 0.61 - 1.42 | 0.743 |

NHIS, National Health Insurance Scheme; NCD, non-communicable disease

## Discussion

This is the largest study on health service utilization among patients with chronic non-communicable disease in Ghana comparing patients in the Ashanti region to the Northern region. The study also reports the impact of reduced HSU on perceived health, evaluated health status and patients’ satisfaction during the COVID-19 pandemic.

We found that there was significant reduction in HSU among patients with NCD in Ghana with a disproportionally significantly reduced HSU in the Northern region (non-epicentre region) than the Ashanti region (epicentre region) though there were increased COVID-19 cases in the Ashanti region than the Northern region. The identified predictors of reduced HSU include decreasing age, Northern region residence, increasing household income, possession of valid health insurance, and increasing duration of NCD.

Over a third of all respondents missed their hospital appointment during the COVID-19 pandemic. This is similar to a study in Ethiopia reporting missed appointment in 26.8% of patients reportedly due to fear of COVID-19 infection and lack of transportation in Ethiopia during the COVID-19 pandemic [26]. In another study in Oman, 23.8% missed their hospital appointments and even higher proportion of 40.5% during the first wave of the pandemic [27]. This finding is remarkable as it had dire consequences on patient care and was higher than a recently published longitudinal study reporting a range of 3.1% - 22.4% in the United States [28]. Missed appointment has been shown to be associated with poor control among patients with diabetic and hypertension in a study in Ethiopia in east Africa [29]. Missing hospital appointment may be associated with patients’ medication shortage and missing opportunities for required laboratory investigations to ensure quality patient care to avoiding complications which may contribute to excess mortality[30].

During the peak of the pandemic, the Government of Ghana instituted lockdown measures to prevent the spread of the disease and this led to reduced HSU which may have led to excess mortality as also shown in a systematic review with 100.3 deaths per 100,000 population per year [30]. NCDs have been documented in several studies as a major risk factor for COVID-19 infections and increased mortality in relation to COVID-19 [11, 31]. It has been argued in several studies that patients with diabetes and hypertension may probably be dying with COVID-19 infection and not necessarily from complications of COVID-19 infection [32, 33] and missed hospital appointment may be a major contributing factor.

Significantly, about a half of respondents in the Northern region reportedly missed their appointment as compared to a fifth in the Ashanti region. Nationally, about a fifth of respondents reported being acutely ill which may have resulted from the missed appointments and inability to access health care. Acute illness was significantly higher in the Northern region (25.1%) than the Ashanti region (11.6%) though there was no significant difference between the admission proportions in the two regions in our study. The difference in response in the regions could be explained by the contextual factors, literacy rate, socio-economic states and excessive fear of COVID-19 in the Northern region due to COVID-19 cases in the hospitals in the Northern region [34]. Hospitalization was reported by about 14% of respondents which may also be as a result of missing of hospital appointment, reduction in HSU and lead to excess mortality [30].

Almost half (43.5%) of all respondents reported that COVID-19 led to a decrease in their HSU. There was significantly a higher proportion reporting decreased HSU in the Northern region 276 (65.4%) than in the Ashanti region 91 (21.6%). This was an unexpected finding considering the fact that there were more cases of COVID-19 infection in the Ashanti Region than the Northern region[19]. Possible reasons to explain the differential reduction include reports of reduced hospital attendance for fear of spread of COVID-19 infection with patients staying away from health facilities in Savelugu in the Northern region of Ghana [34], which was similarly reported among patients in Ethiopia [35]. The reported reduction in HSU in our study was lower as compared to a single centre study in Ghana where there was a 50% reduction in OPD attendance observed at a university hospital [36]. A scoping review including 262 articles, generally reported a median reduction in HSU of 36% as a result of the COVID-19 pandemic with other studies also reporting reduced HSU [9, 37, 38].

In a study in Ghana by Fenny et al, there was a reduction in health insurance claims by 20% but an increase in complication by 18.6% among patients with hypertension [20]. These disruptions in hypertension and diabetes management is not isolated to Ghana. Hypertension clinics have been reported to be disrupted in 59% of countries and diabetes management disrupted in 56% of countries during the first quarter of the year 2020 during the COVID-19 pandemic [39]. Another study in Ghana by Sarfo et al[40], reported an increase in NCD-related complications such as stroke in a tertiary institution with increase in case fatality rate of 29.3% in 2020 from 24.2% in 2019 though seasonal variations were not accounted for in their study.

Of all the predisposing factors, enabling and need factors proposed by the Anderson’s Behavioural Models of HSU [15–17], decreasing age and Northern region residence were identified as predisposing factors predicting reduced HSU. Increasing household income and possession of a valid health insurance were identified as the enabling factors predicting reduced HSU in Ghana as a result of the COVID-19 pandemic. Increasing duration of illness was the only need factor that predicted decrease in HSU during the COVID-19 pandemic.

Increasing age has been reported as a predictor of increased HSU in several studies [41–43]. This may be because older patients have multiple co-morbidities warranting frequent HSU for reviews, admissions and laboratory workup as shown in other studies [41, 44]. They were also at risk of COVID-19 infections and increased COVID-19 related mortality [45]. Those with hypertension, diabetes, heart diseases were at increased risk of severe COVID-19 disease and increased risk of death [46]. Another study in Korea reported a decreased HSU with increasing age [47] as most may resort to over-the-counter medications, herbal medications or get used to their medical conditions and hence would not report to formally to a health facility. Other studies reported equivocal findings with respect to increasing age and increased HSU [48, 49].

We also found HSU to be decreased significantly in the Northern region than the Ashanti region. The predominant ethnicity in the Northern region is Dagomba and the Akans dominate the Ashanti region. The Northern region was a significant predictor of decreased HSU. This reflects the increased fear among inhabitants of the Northern region as compared to the Ashanti region even when the number of cases in Ashanti region were significantly higher than the Northern region [19, 50]. This is the most striking finding in our study. Though COVID-19 infection had a high incidence rate in the Ashanti region hence noted as an epicentre region there was more reduction in HSU in the Northern region. Total number of COVID-19 cases were 22,640 in the Ashanti region as compared to just 1,930 in the Northern region as at April 2024 [19]. This could be explained by the perception, response and health seeking behaviour of the people. The level of knowledge could have impacted on their reduced HSU worsened by the fear of COVID-19 infection especially when a hospital in Savelugu in the Northern region was found to have high number of staff contracting the COVID-19 infection [34]. This phenomenon led to significantly more patients reporting of missing hospital appointments in the Northern than Ashanti region and more reported acute illness in the Northern region than the Ashanti region (25.1% vs 11.6%) in our study.

High household income levels were associated with increased odds of reduced HSU with aOR of 1.36 (CI-1.15-1.60), p<0.001). According to Anderson’s Behavioural theory of HSU [16], income is an enabling factor for increased HSU. There is evidence from some studies to suggest that people with low socio-economic status just above the poverty line are less likely to use health service [51, 52]. People with high socio-economic status tend to use private medical and specialist services than those of low socio-economic status [53]. People with high income are able to access health service when required and may have access to telemedicine and may not report to health facility. Studies have also shown that those with high income are likely to live well and not be predisposed to health conditions that would increase their HSU. Income level determines the level of engagement with the health facility. Those with low socio-economic status are more likely to visit the general practitioners and emergency care while those with high socio-economic status are more likely to visit specialists [43, 48].

We also found that possession of valid national health insurance predicted significantly decreased HSU (aOR=7.42 95% CI 2.44 - 22.57, p< 0.001). Possession of a valid health insurance has been described by Andersen et al [16] as an enabling factor for HSU. Most studies have shown that HSU increases with ability to access health care and the possession of valid health insurance in South Africa [54], in Mongolia [42] in Iran [55], in Ecuador [56] and Mexico [57]. Our study findings were contrary to these global studies reporting increase in HSU with possession of valid health insurance. Though there was high percentage access to insurance in Ghana, it did not improve HSU during the COVID-19 pandemic but rather was associated with decreased HSU. This may have been as a result of the fact that those with valid health insurance decided not to use the health facility for fear of contracting the COVID-19 infection, were rescheduled for longer review dates or may have considered alternate ways to seek health care including private facilities as the major users of health service before the pandemic. A previous study in Ghana before the COVID-19 pandemic showed that enrolling onto health insurance increases the likelihood of prescriptions, clinic visits and HSU [58]. Those who utilize health service were more likely to renew their health insurance than those who do not utilize [59]. Though it is generally accepted that increased enrolment on the national health insurance scheme is associated with increased HSU this was not shown among the elderly poor especially in living in rural areas in Ghana [60]. Other plausible reasons for this observation is the fact that those who possess the health insurance decreased their HSU in the public facility because that is where the insurance card is accepted and hence the those who have the card will therefore not utilize the health facility due to the cancelation of out-patient’s clinics as shown in another study [27] or fear of contracting COVID-19 infection the health facilities as observed in Savelugu in the Northern region [34]. Such patients may then seek care elsewhere or wait until their new scheduled visit was communicated after the lockdown.

We found that longer duration of the NCD, was associated with reduction in HSU (aOR=1.03, CI1.01-1.06, p=0.033). Several studies have shown a linear relationship between duration of chronic illness and HSU [42, 47, 61, 62]. Patients with chronic conditions like NCDs have been shown to have an increased HSU but in the light of COVD-19 as shown in our study, there was rather a decrease in HSU during the COVID-19 [43]. This may be as a result of the fact that most clinicians gave longer review dates to patients with stable chronic NCDs and hence reduction in HSU during the COVID-19 pandemic as found in our study. Those with longer duration may also be very experienced in the management of their disease and hence were not necessarily at risk of poor control to come for review except when complicated. On the contrary, those with shorter duration may need shorter reviews and may not be experienced in managing their cases at home during the lockdown period. Those with longer duration of NCDs were more likely to be older and hence may consider themselves at increased risk of severe COVID-19 and COVID-19 related mortalities and hence would stay away from the health facilities.

Our study had some limitations. This was a hospital-based study and may have missed responses from those who do not use formal health facilities. The study was restricted to public health facilities and may have missed valuable information from patients in private facilities and those using alternative medicines.

### Recommendations

We recommend further prospective and qualitative studies in pandemic situations to better understand the contextual factors that may account for the reduced HSU in the Northern than the Ashanti region. Effort should be made by the Ghana Health Service to address geographical and financial access barriers with mobile testing, and emergency transport services so people get the care they need especially in pandemic situations. Health care managers should provide PPEs, adequate staff training on infection prevention and evidence-based scheduling of appointment to reduce spread of infection. They should ensure HSU for NCDs are not affected in pandemic situations at they are at increased risk of infection and mortality. There is also the need to intensify public education by health institutions to allay fears and encourage HSU when needed as fear of contracting COVID-19 infection was a major reason for the decreased HSU in Ghana.

## Conclusion

Despite a relatively lower COVID-19 burden in Ghana, there was significant reduction in HSU, with greater reduction in HSU in the Northern Region (a non-epicentre) than the Ashanti region (an epicentre) though there were more cases of COVID-19 infection in the Ashanti region than the Northern region. The predictors of reduced HSU were found to be increasing age, residency in the Northern region, increasing income level, possession of valid National health insurance and increasing duration of NCD in our study. This calls for more attention in non-epicentres and in patients with NCDs in future pandemics.

## Data Availability

All data generated or analyzed during this study are included in this published article

## Acknowledgement

I will like to acknowledge Ghana West African Centre for Global Health and Pandemic Prevention (G-WAC) for funding my PhD work through the DAAD and all the hardworking staff of G-WAC.

## Authors contributions

EKT initiated the study and put together the initial draft with ideas and inputs from WQ, RB, JA and SN. SG, JS, RA were involved in the collection and cleaning of the data. Data analysis was done by EKT. Figures were designed by EKT after the initial concepts were drafted by EKT. WQ and SN contributed to the write-up of the discussion in the manuscript. All authors read and approved the final manuscript.

## Funding

This study was part of a PhD program funded by the Ghana West African Centre for Global Health and Pandemic Prevention (G-WAC) through the DAAD (German Academic Exchange Service) scholarship but there was no funding for publication.

## Availability of data and materials

All data generated or analyzed during this study are included in this published article

## Declarations

### Ethical approval and consent to participate

Ethical clearance was granted from the Ghana Health Service Ethics Review Committee with approval number GHS-ERC/010/01/24, the Committee on Human Research and Publication Ethics (CHRPE) of KNUST with approval number CHRPE/AP/1045/23 and the institutional review board of the Komfo Anokye Teaching hospital with approval number KATH/IRB/AP/009/24 before the conduct of the study. We confirm that all methods were carried out in accordance with the Declaration of Helsinki.

### Consent for publication

Not applicable.

### Competing interests

The authors declare no competing interests.

## Notes

### Competing Interest Statement

The authors have declared no competing interest.

